# Accurate and reproducible prediction of ICU readmissions

**DOI:** 10.1101/2019.12.26.19015909

**Authors:** Dinh-Phong Nguyen

## Abstract

Readmission in the intensive care unit (ICU) is associated with poor clinical outcomes and high costs. Traditional scoring methods to help clinicians deciding whether a patient is ready for discharge have failed to meet expectations, paving the way for machine learning based approaches. Freely available datasets such as MIMIC-III have served as benchmarking media to compare such tools. We used the OMOP-CDM version of MIMIC-III (MIMIC-OMOP) to train and evaluate a lightweight tree boosting method to predict readmission in ICU at different time points after discharge (3, 7 and 30 days), outperforming existing solutions with an AUROC of 0.805 for 3-days readmission.

## Introduction

Recent studies have shown that readmission in the intensive care unit (ICU) is associated with poor clinical outcomes, increased length of ICU and hospital stay, and high costs (1, 2). One of the main reasons for ICU readmission that has been identified is premature discharge (3); in fact the transfer of patients from an ICU to a general hospital ward represents a high-risk event, and thus the decisions about which patients are ready to be discharged are daily struggles for ICU clinicians (4). Other studies have shown that determining the best timing for ICU discharge is usually based on subjective intuitions and that readmission prediction tools can help physicians in this endeavor, provided their performance and ease of adoption (5, 6). As traditional scores based on logistic regression or Cox proportional hazards models such as the Stability and Workload Index for Transfer score (SWIFT) or the LACE index have failed to meet expectations (6–10), numerous prediction models using machine learning have been proposed in the recent past, several of which trained and evaluated on the Multiparameter Intelligent Monitoring in Intensive Care (MIMIC-II or MIMIC-III) open database (11–15).

MIMIC-III is a large ICU EHR database widely accessible to researchers internationally under a data use agreement, allowing clinical studies to be reproduced and benchmarked (16, 17). In order to make multicenter ressearch possible, a valuable effort has been made to convert MIMIC-III to the Observational Medical Outcomes Partnership common data model (OMOP-CDM), which provides structural and conceptual models relying on international reference terminologies (18, 19). For the sake of reproducibility and ease of subsequent implementation in other centers using the same data model, we chose to use the OMOP-CDM version of MIMIC-III (MIMIC-OMOP), for which documentation and a mapping Extract-Transform-Load (ETL) process are freely available (20).

### Related works

Among the numerous works aiming to provide decision-making tools for ICU clinicians at discharge time, two in particular caught our attention in terms of performance and similarity of setting to our own.

Lin et al. (12) proposed an advanced neural network for 30-day ICU readmission prediction (LSTM-CNN based model) achieving an Area Under Curve of the Receiver Operating Characteristic (AUROC) metric of 0.791 on MIMIC-III, using chart events 48h time series, diagnostic ICD-9 codes embeddings, and demographic information of the patients. The authors claim to offer higher sensitivity (0.742) compared to existing solutions, regardless of the specificity trade-off. With a fixed specificity at 0.850 and 0.800, they achieve a sensitivity of 0.548 and 0.619 respectively with their best model. There is no mention of precision nor F1-score. Pakbin et al. (13) trained a simpler and more interpretable gradient boosting model (XGBoost) for predicting risk of ICU bounceback and readmission at a variety of time points using MIMIC-III, achieving AUROC of 0.76 and 0.75, F1-score of 0.20 and 0.34, for 72h and 30-days ICU readmission respectively. They use chart events time series, ICD-9 codes indicators, as well as admission, demographic and length-of-stay information of the patients. There is no mention of sensitivity, specificity or precision.

## Methods

### Data and patients

MIMIC-III integrates deidentified, comprehensive clinical data of patients admitted from 2001 to 2012 at the Beth Israel Deaconess Medical Center in Boston, Massachusetts. We restricted our analyses to ICU stays of patients over 18 years old, ending up with a dataset of 55135 stays. Variables used in the model were age, gender, length of stay, provenance from a surgery ward, current count of ICU visits for the same patient (=visit rank), and three values for a number of measures and blood tests: the first entry for a given stay, the last one and the absolute difference between the two. Those measures were total glasgow coma scale (GCS), motor GCS, verbal GCS, eye movement GCS, systolic blood pressure, heart rate, respiratory rate, body temperature, oxygen saturation, oxygen inspired fraction, body weight, urine output, serum bicarbonate, serum urea, total bilirubin, serum sodium, serum potassium, serum creatinine, blood platelets, hemoglobinemia, blood hematocrit, blood leukocytes, serum lactates, blood PH, blood glucose and the International Normalized Ratio (INR). Three variables were extracted from medical and nurses text notes: history of AIDS, metastatic cancer and/or of advanced hematologic condition (myeloma, lymphoma or leukemia). Finally, one feature counting the number of available values for all the previous variables was added. Missing data were then imputed via multivariate imputation from all other available variables (21), with a Bayesian ridge regression as the estimator (22). A short summary of the dataset’s main characteristics is reported in Table1.

**Table 1.** Description of the dataset’s main characteristics

|  |  |  |
| --- | --- | --- |
| Total ICU visits, n |  | Overall<br>55135 |
| Gender, n (%) | Male | 31219 (56.6) |
|  | Female | 23916 (43.4) |
| Length of stay (days), mean (SD) |  | 3.2 (4.9) |
| Age, n (%) | 18-25 | 1346 (2.4) |
|  | 25-45 | 6271 (11.4) |
|  | 45-65 | 18887 (34.3) |
|  | 65-85 | 22800 (41.4) |
|  | 85-95 | 3226 (5.9) |
|  | 95+ | 2605 (4.7) |
| Visit rank, mean (SD) |  | 0.2 (0.5) |
| Provenance, n (%) | Other wards | 27391 (49.7) |
|  | Surgery | 27744 (50.3) |
| Personal history of AIDS, n (%) | No | 53013 (96.2) |
|  | Yes | 2122 (3.8) |
| Personal history of metastatic cancer, n (%) | No | 50076 (90.8) |
|  | Yes | 5059 (9.2) |
| Personal history of advanced hematologic condition, n (%) | No | 52359 (95.0) |
|  | Yes | 2776 (5.0) |

### Outcome definition

We used a similar outcome definition to Lin et al. (12), where positive cases were regarded as the visits where the patients could benefit from a prediction of readmission before being transferred or discharged: visits where the patients were either transferred or discharged but returned to ICU, or died before a defined time limit (3, 7, or 30 days).

### Model training and evaluation

Several model families were tested in a screening phase, among which linear models, support vector machines, naive bayes, decision trees and ensemble methods. XGBoost, a gradient tree boosting method that is widely used to achieve state-of-the-art results on many machine learning problems, was consistently outperforming the others on all metrics and was thus selected as the prediction model for this study (23). Schematically, gradient boosting methods work by iteratively fitting “weak” models to the residuals of the previous model, and adding the newly estimated residuals to the previous model’s prediction, thus forming a “new” prediction, and so on until a stopping criterion is met. XGBoost implements this algorithm with decision trees, an additional custom regularization term in the objective function, and several computing tweaks to optimize speed and performance, such as parallel learning or sparsity awareness.

Our dataset was divided into three parts, each representing 64%, 16% and 20% of the whole respectively: the first part to train our model (the *training set*), the second to tune the model’s output classification threshold and to apply the *early stopping* method (the *validation set*) and the third part to evaluate the performance of our model on unseen data (the *test set*). The splits were stratified and grouped by individual patients, meaning each set contained about the same proportion of each outcome class and that all visits of a patient were grouped in the same set.

To reduce overfitting, we used the *early stopping* method. At each training epochs, the model’s performance is evaluated on the validation set by measuring the negative log-likelihood: if it hasn’t improved after a fixed number of epochs (in our case 10), then the training is stopped. As for the final evaluation, we decided to report a comprehensive set of metrics, for a full overview of the model’s abilities and to ease future comparability with other approaches: area under the receiver operating characteristic curve (AUROC), precision (positive predicted value), specificity (true negative rate), sensitivity (true positive rate, also known as the recall) and F1-score, the harmonic mean between precision and recall. All metrics were calculated over the test set.

The probability threshold above which the output of our model would be classified as a positive outcome has been chosen according to an iterative procedure optimizing for the highest F-*β* score on the validation set, with *β* = 1.5. For reference, the F-*β* score is a weighted weighted harmonic mean between precision and recall, favoring recall when *β >* 1, and vice versa. Hyperparameter selection was done via a stratified group 5-fold cross-validation procedure in a grid-search setting, optimizing for the following subset of parameters: the number of trees, the maximum depth for each tree, the proportion of features used for each tree and the learning rate. Calibration of the models was assessed by separating the predicted probabilities over the test set into deciles, and assessing the proportion of realized outcomes in each bin. A model is said to be well calibrated when each bin’s true outcome proportion is close to the bin’s predicted probabilities, resulting in a calibration curve close to the diagonal line when plotted.

Apart from performance, we also wanted to understand which features were important to the model. Contributions of each feature were reported using the *TreeExplainer* from the SHAP python library, a state-of-the-art explanation framework for tree based-methods that enables the tractable computation of optimal local (ie. per sample) explanations, as defined by desirable properties from game theory elements, such as Shapley values (24, 25). It is important to note that this might not necessarily mean that features with a higher importance are significantly associated with the outcome in a causal relationship, but it is nonetheless the best available way to assess the internal correlations learned by the model.

## Reproducibility

All code used to produce this work, essentially written in Python, is available at http://github.com/deepphong/icu-readmission, accompanied by a step by step example Jupyter notebook and generic functions to exploit any database in OMOP format for other use cases.

## Results

The evaluation results on the test set are reported in Table2 for each metric and outcome. Overall, at the classification threshold set for optimal F1.5-score, our model consistently equals or outperforms the results previously reported by Lin et al. (12) for 30-days readmission (AUROC 0.794 vs. 0.791, recall 0.796 vs. 0.742) and Pakbin et al. (13) for 3-days, 7-days and 30-days readmission (AUROC 0.805 vs. 0.76, 0.807 vs. 0.77 and 0.794 vs. 0.75 respectively; F1-score 0.481 vs. 0.22, 0.527 vs. 0.32 and 0.560 vs. 0.37 respectively). Performance for other classification thresholds are reported in Figure 1 for 3-days readmission prediction.

**Fig. 1.**
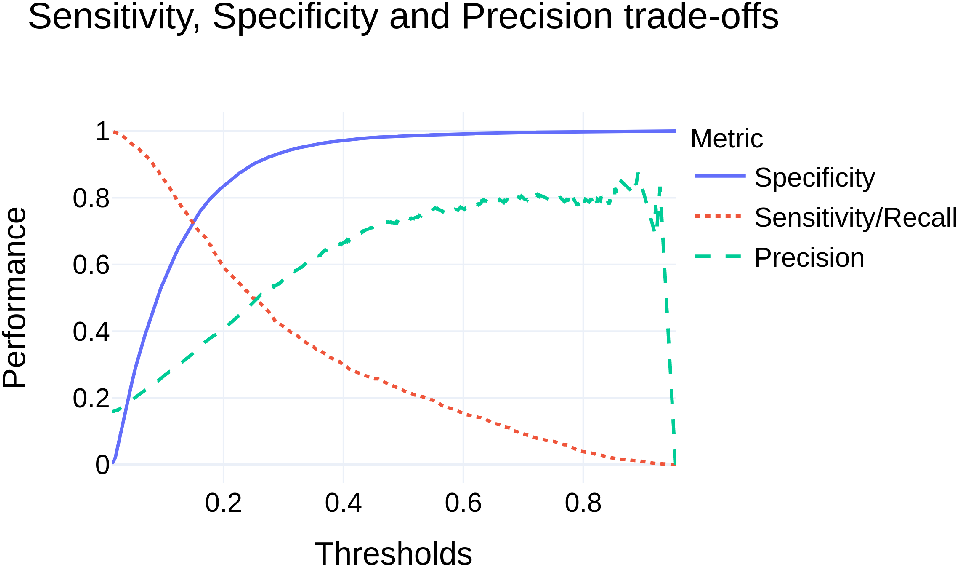
Performance trade-offs for specificity, sensitivity and precision on the test set according to different classification thresholds, on 3-days readmission prediction

**Table 2.** Performance metrics over the test set for the three different outcomes, at the classification threshold for optimal F1.5-score. CIR = class imbalance ratio (proportion of positive cases among the population)

|  | 3-days readmission<br>(CIR=15.8%) | 7-days readmission<br>(CIR=19.7%) | 30-days readmission<br>(CIR=27.3%) |
| --- | --- | --- | --- |
| AUROC | 0.805 | 0.807 | 0.794 |
| Specificity | 0.812 | 0.757 | 0.604 |
| Recall/sensitivity | 0.633 | 0.725 | 0.796 |
| Precision | 0.388 | 0.418 | 0.432 |
| F1-score | 0.481 | 0.527 | 0.560 |

Calibration of the model was overall very good, as visually assessed in Figure 2, with a near perfect fit towards the extremes, meaning that the model is more frequently right the more confident it is in making its predictions. While calibration statistical tests exist, such as the Hosmer-Lemeshow test, we feel like a calibration plot gives more information and intuition.

**Fig. 2.**
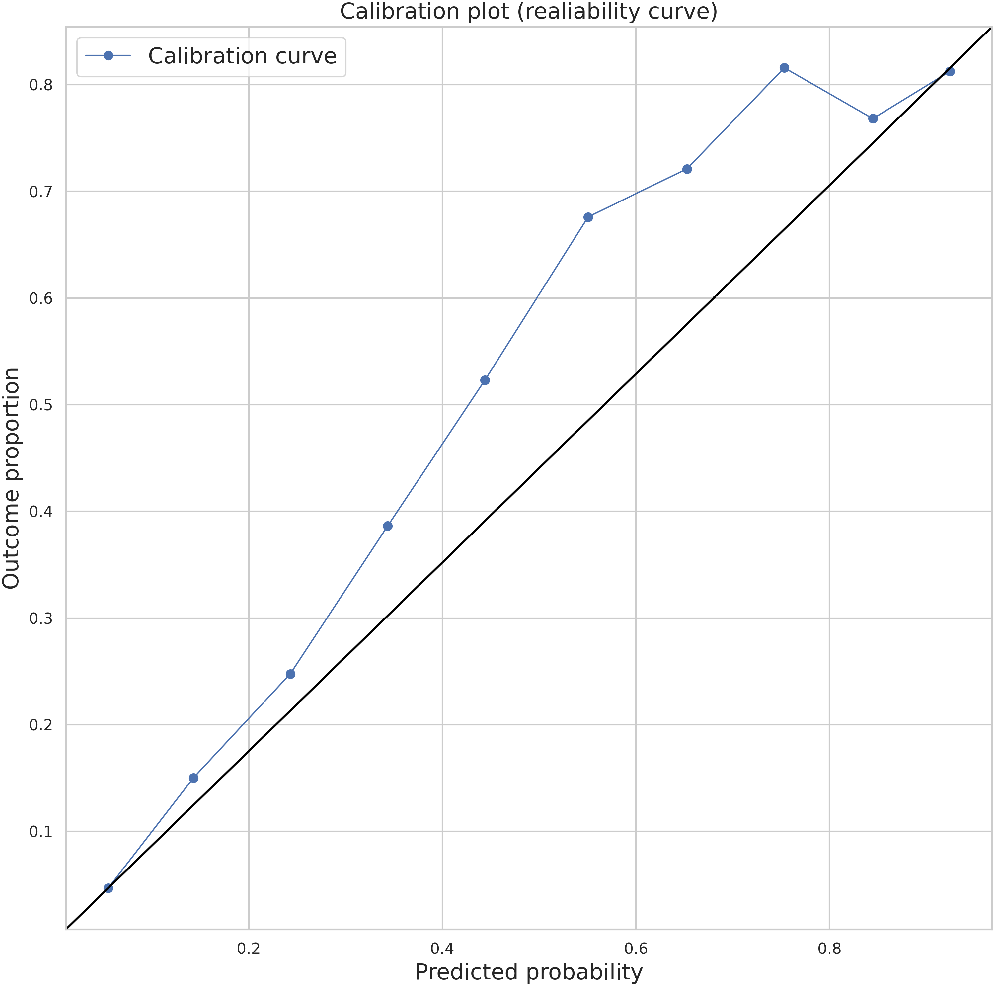
Calibration plot showing the actual proportion of readmission in each decile of predicted probability of readmission for 3-days readmission predicsion

Figure 3 shows the model’s top 20 features importance for 3-days readmission prediction, sorted by the sum of Shapley value magnitudes over all samples. While the length-of-stay seems to be the most discriminative feature for model, with shorter duration associated with a higher chance of readmission, an interesting observation is the feature that comes just after: the number of non-missing features of an individual for a given stay. This feature can be assimilated to the number of different measurements and blood tests taken on an individual for a given stay, and is plausibly correlated with the severity of the patient’s condition.

**Fig. 3.**
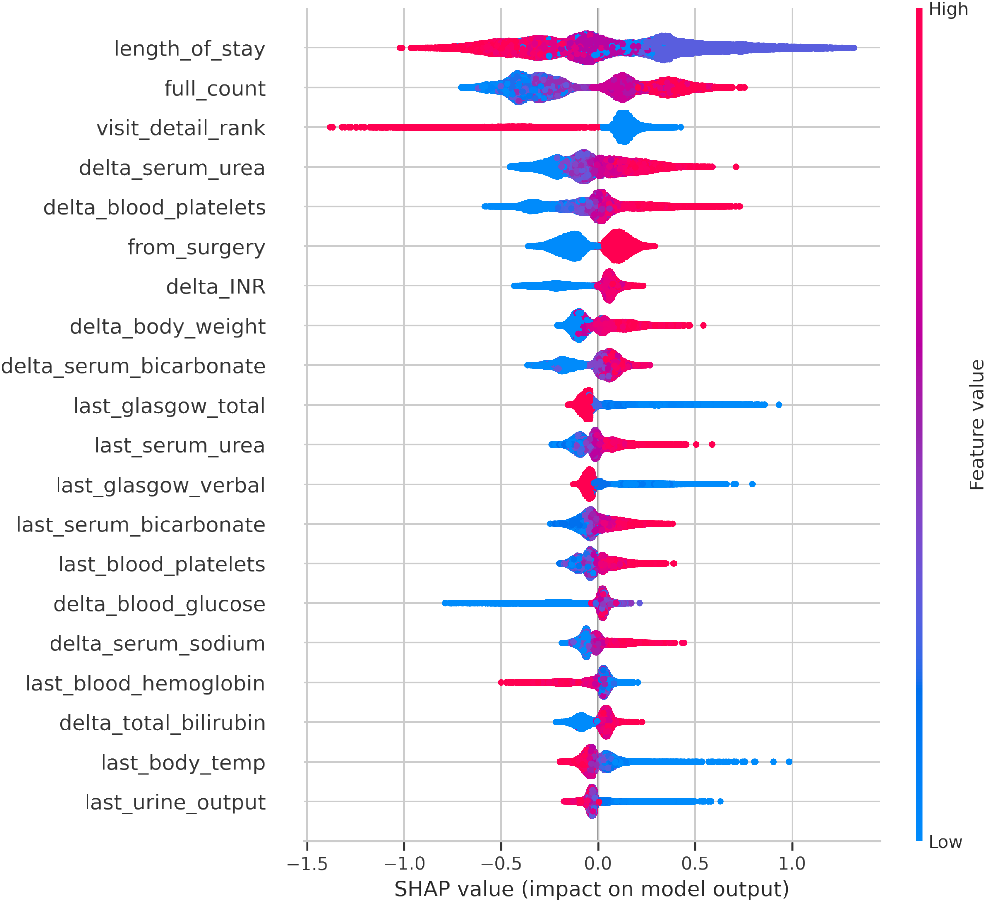
*TreeExplainer* features importance (top 20) on the test set, ranked by the sum of Shapley value magnitudes over all samples for the model prediction. Each dot in the visualization represents one prediction. The color is related to the real data point; if the actual value in the dataset was high, the dot is colored in red; blue indicates the actual value is low. Features prefixed with “last_” are the last available measurements before discharge and features prefixed with “delta_” are the difference between the first and last available measurement of the stay

Performance trade-offs, calibration plots and features importance plots for the other evaluated outcomes are available in the appendix.

## Conclusion and discussion

In this work, we proposed a model based on a tree boosting method to predict ICU readmission at 3, 7 and 30 days using data of from the patients visit available at discharge on the freely available MIMIC-III database. Our solution is open-source and has the advantage of having been conceived with the OMOP-CDM standard, allowing for easier external validation and implementation. While this work improves on existing solutions for ICU readmission prediction, several points still need to be addressed.

The prediction model was trained and evaluated on MIMIC-OMOP, and as of this time, no external validation has been conducted yet. To facilitate this process, all code needed to reproduce the results has been open-sourced. Although efforts have been put into its ease-of-transfer on any other electronic health records (EHR) database using the OMOP-CDM standard, some non neglectible amount of work is always needed to adapt the code to new data. The feasibility of external validation is currently being evaluated on the largest French EHR database. Further work also needs to be done to integrate and evaluate such prediction models in clinical practice. The choice of developing on common health data standards such as OMOP-CDM and/or HL7’s Fast Health Interoperability Resources (FHIR) (26) is a step forward in this direction, as more and more applications are being made compatible with those.

Although there are numerous similar studies claiming state-of-the-art performance for machine learning models on various tasks, most only report one or two metrics, mainly AU-ROC. We argue that it is only by reporting a fully comprehensive set of metrics that models can be made comparable and reproducible. The strengths and weaknesses of a model often rely on the performance trade-offs; depending on the use-case, one would want to favor one metric over the others (eg. sensitivity for non-invasive cancer screening), and would be able to assess the model’s ability to do so with reports such as Figure 1.

Apart from our model’s intrinsic performance, another interesting finding was the importance of including a variable accounting for the available measures among the ones selected for the model. This shows that the missing measures were missing not at random (MNAR), and the rationale behind this seems to be that patients with a poorer prognosis usually have more tests and measurements done to them. This feature could possibly be indirectly correlated with the care providers’ overall feeling of the patient’s current state, and we postulate that existing models could be improved by adding similar information.

## Data Availability

All data used and code are freely accessible.

https://mimic.physionet.org/

https://github.com/deepphong/icu-readmission

## Declarations

### Ethics approval and consent to participate

Not applicable.

### Consent for publication

Not applicable.

## Acknowledgements

This work has been made possible thanks to the support of the WIND-DSI department of AP-HP hospitals and the DIM department of Bicetre hospital, especially Christel DANIEL and Marie FRANK. Special thanks to Julien DUBIEL from WIND-DSI for his constant help with logistics.

