## Appendix for "Accurate and reproducible prediction of ICU readmissions"

### Appendix 1: Supplementary figures

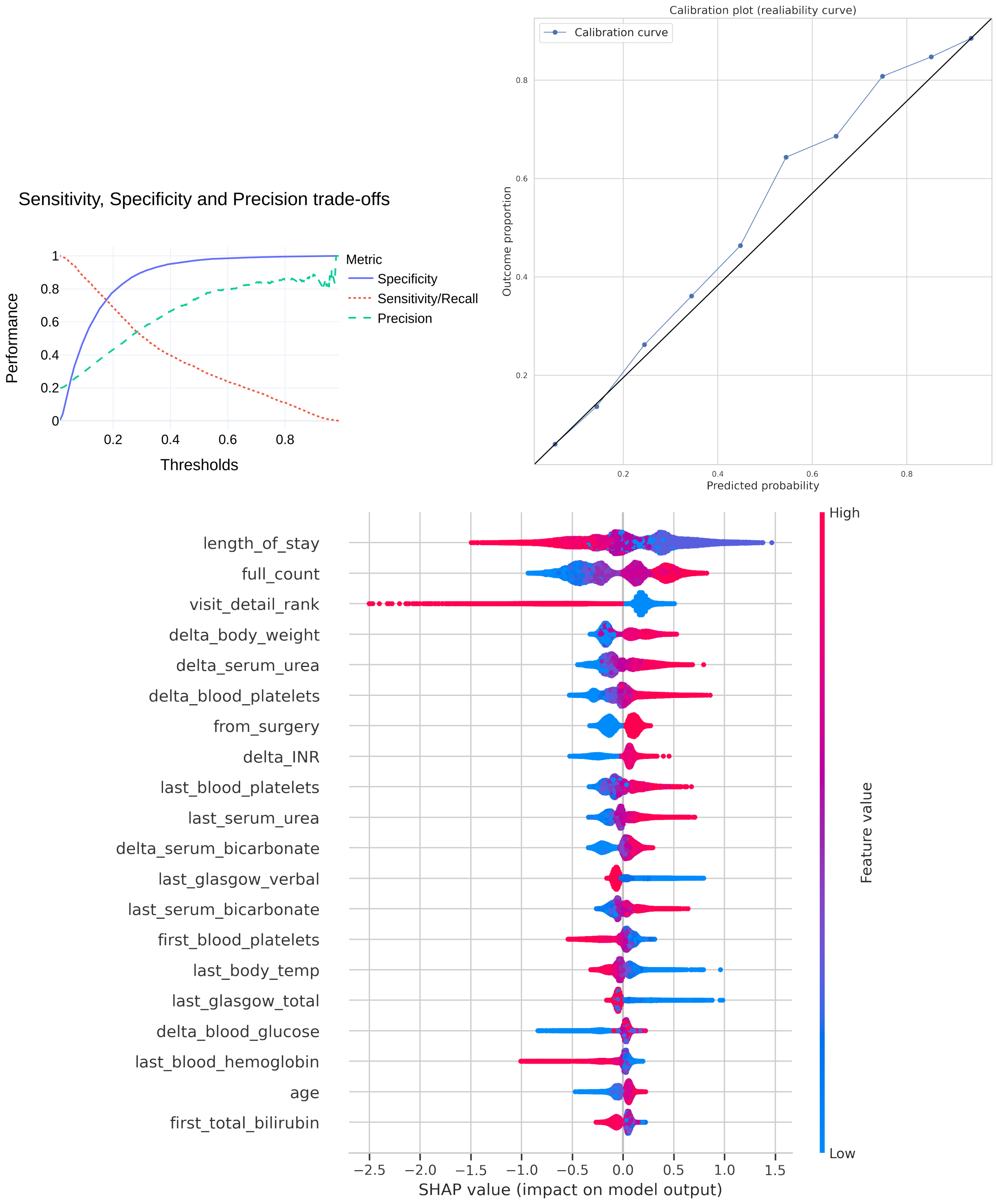


**Fig. 4.** Performance trade-offs, calibration plot and features importance plot for 7-days readmission prediction

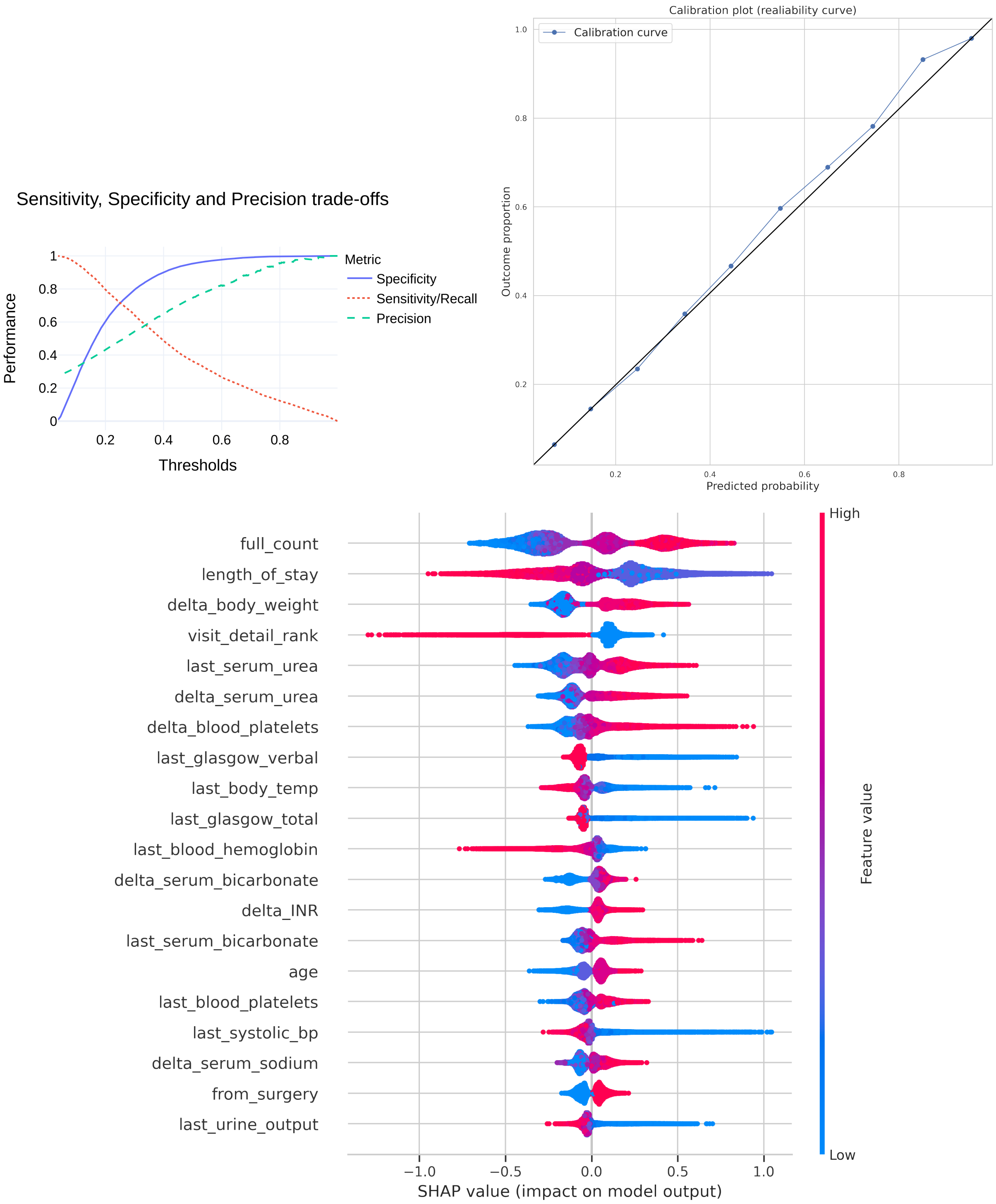


**Fig. 5.** Performance trade-offs, calibration plot and features importance plot for 30-days readmission prediction
